# Long-Term Effectiveness of a Digital Therapeutic Intervention for Smoking Cessation: A Randomized Controlled Trial

**DOI:** 10.1101/2021.08.19.21262270

**Authors:** Jamie Webb, Sarrah Peerbux, Alfonso Ang, Sarim Siddiq, Yusuf Sherwani, Maroof Ahmed, Hannah MacRae, Hannah Puri, Azeem Majeed, Suzette Glasner

## Abstract

**Introduction:** The present study evaluated the long-term effectiveness of Quit Genius (QG), an extended digital smoking cessation intervention.

**Methods:** Participants were adult smokers (N=556) recruited between January and November of 2019 via social media and referrals from primary care practices who were given nicotine replacement therapy and randomly assigned to Quit Genius (QG) (n=277), a cognitive behavioral therapy (CBT) based digital intervention or Very Brief Advice (VBA) (n=279), a face-to-face control intervention. Primary analyses (N=530), by intention-to-treat, compared QG and VBA on biochemically verified continuous 7-day abstinence at 4, 26, and 52 weeks post-quit date. Secondary outcomes included sustained abstinence, quit attempts, self-efficacy and mental well-being.

**Results:** Seven-day point prevalence abstinence from weeks 4 through 52 ranged from 27% to nearly 45% among those who received QG, and from 13% to 29% for those in VBA. Continuous 7-day abstinence at 26 and 52 weeks occurred in 27.2% and 22.6% of QG participants, respectively, relative to 16.6% and 13.2% of VBA participants; QG participants were more likely to remain abstinent than those in VBA (Relative Risk [RR]= 1.71, 95% CI 1.17-2.50; *p*=0.005).

**Conclusions:** This study provides evidence for the long-term effectiveness of an extended digital therapeutic intervention.

**Implications:** The long-term effectiveness of digital smoking cessation interventions has not been well-studied. This study established the long-term effectiveness of an extended CBT-based intervention; results may inform implementation of scalable, cost-effective approaches to smoking cessation in the health system.

## Introduction

Smoking is the leading cause of preventable death worldwide, causing over 8 million deaths annually ^1^ with a total annual global economic cost exceeding $1.4 trillion ^2^. Despite advances in pharmacological and behavioral smoking cessation treatment approaches, even when evidence-based treatments are combined with best practice guidelines, long-term abstinence rates remain relatively low, frequently falling below 20% at 1 year post-intervention ^3,4^. One possible explanation for the lack of longer-term success is that the most commonly used treatment models do not align well with the scientific understanding of addiction as a chronic illness. Given that tobacco addiction is characterized by a chronic and relapsing clinical course, extended treatment models that support multiple transitions between relapse and recovery may optimize outcomes ^5^. Though traditional smoking cessation support involves low intensity motivational advice, studies combining pharmacological smoking cessation treatment with extended, evidence-based counselling ranging from 6 ^5,6^ to 12 months in duration ^7^ have produced superior clinical outcomes, both in terms of relapse prevention and abstinence rates. Nevertheless, utilization rates of evidence-based smoking cessation treatment programs are astonishingly low, with as few as 5% of smokers accessing care ^8^. Moreover, given the limited scalability of face-to-face smoking cessation treatment models, novel approaches to broadening the reach of evidence-based treatments are urgently needed.

Progress in mobile technology has led to the proliferation of mobile health (mHealth) approaches to smoking cessation, conferring many advantages over face-to-face approaches, including low cost, greater accessibility, customizable features to support individuals through different psychological stages of change, and scalability. Nevertheless, scientific support for the relatively large existing base of mHealth tobacco cessation interventions is lacking. According to a recent systematic review, among the 50 most highly recommended apps suggested by leading app stores, only 4% had any scientific support ^9^, and the majority of existing mHealth apps underutilize evidence-based treatment strategies.

Following evidence-based treatment guidelines, our group developed Quit Genius (QG), a digital therapeutic intervention combining pharmacotherapy with cognitive behavioral therapy (CBT) within an extended treatment model. QG is a 52-week cessation program combining digital CBT with coaching support delivered asynchronously within the app. Concurrent access to nicotine replacement therapy (NRT) is provided to each participant. We recently reported promising preliminary short-term outcomes of QG, with a CO-verified 4-week quit rate of 44.5% ^10^. However, the extent to which these short-term outcomes are sustained over an extended follow up period remains unknown. As such, in the present study, we examine the effect of QG on longer-term outcomes through 52 weeks post-quit date. To achieve this, we evaluated continuous smoking abstinence at 4-, 26- and 52 weeks post-quit date (QD). The impact of QG, relative to the control condition on secondary outcomes including quit attempts, psychological well-being, and self-efficacy was also examined.

## Methods

### Study Design and Participants

We conducted a single-blinded, two-arm parallel design, randomized controlled trial (1:1 allocation ratio), with 4-, 26- and 52-week follow-up. The trial was registered in the International Standard Randomized Controlled Trial Number (ISRCTN) database (https://www.isrctn.com/ISRCTN65853476) on December 18, 2018. This study complies with the Declaration of Helsinki and ethical approval was granted by the Health and Social Care Research Ethics Committee A (HSC REC A; reference 18/NI/0171).

Participants were recruited via social media and referrals from primary care practices in the United Kingdom between January and November 2019. Participants were directed to a study website, where they were provided with study information and prompted to complete pre-screening questions. They were then invited to an in-person baseline assessment in which eligibility was confirmed. Subsequently, informed consent was obtained and baseline data were collected.

Individuals were eligible to participate if they were adult smokers (aged ≥18 years), smoked ≥5 cigarettes a day for the past year, had the required mobile phone functionality (≥5^th^ generation iPhone or version ≥18 Android) and were not using any other form of behavioral or pharmacological support for smoking cessation. Exclusion criteria included not speaking English, pregnancy, COPD, currently taking psychiatric medication, and any serious health conditions that would hinder completion of the study procedures. A total of 556 participants consented to participate, completed screening, were offered NRT and randomly assigned to either QG or the control condition.

### Randomization and Blinding

Participants were randomized with an allocation ratio of 1:1 (treatment:control) using a block size of 4 participants. An online research management system (curebase) was used, with researchers blinded to treatment allocation until after randomization had been performed.

### Treatment Intervention: Quit Genius

QG ^11^ is a 52-week intervention informed by the principles of CBT. QG comprises a smartphone application with self-guided CBT content, coupled with a quit coach who provides asynchronous messaging to reinforce the use of CBT skills. QG utilizes components that have demonstrated efficacy in promoting smoking cessation, including encouraging medication adherence, progress feedback, goal setting and self-monitoring. The application collects user data that help personalize the pace, feedback, and content each participant experiences during the program. The variables that inform personalization include app utilization, program completion, motivations for quitting smoking, reasons for continuing to smoke, and quit-date.

Tailored content is delivered in the form of animated videos, audio sessions, reflective exercises, and quizzes. The user is prompted to complete a series of consecutive self-paced steps, based on principles of motivational enhancement therapy and CBT, on their smoking cessation journey. QG’s content is divided into two stages: ‘Essentials’ and ‘Sustain.’ In the Essentials stage, the user is prompted to complete a series of digitally delivered psychoeducation and motivational exercises, including preparing for the quit date, review of the research evidence supporting use of NRT as part of cessation treatment, how NRT works, how to use it, and thinking about their personal reasons for quitting. In the ‘Sustain’ stage, post-QD, the focus is on relapse prevention, supporting long-term abstinence. Users are encouraged to monitor their smoking habits daily by logging the number of cigarettes smoked, their smoking triggers, and the intensity of their cravings. After quitting smoking, participants are encouraged to log their smoking status when they open the app. Accordingly, participants receive tailored feedback concerning abstinence status and associated health benefits over the course of their involvement in the program.

QG participants had access to a ‘Quit Coach’, an advisor qualified by the National Centre for Smoking Cessation and Training. The coach provides personalized CBT and motivational support via the in-app chat and phone. At baseline, participants are scheduled for a 30 minute phone call with their Quit Coach. During this call, participants receive an introduction to QG, discussion of their individualized quit plan, and guidance concerning the availability, types, and methods of using NRT. Subsequently, interactions with the quit coach transition to the in-app chat platform. Features include: (1) the ability for individuals to monitor their progress via the app, which details health improvements and financial benefits gained from becoming and remaining abstinent, and (2) access to a ‘Craving Toolbox’ comprising audio delivered mindfulness, meditation and breathing exercises designed to help the user manage cravings.

The QG app also provides CBT based skills training addressing mental health concerns commonly linked with tobacco use, including anxiety, mood, stress, self-esteem and social skills, as well as general health and well-being practices such as diet, exercise, and self-care. CBT-based techniques include goal setting, cognitive restructuring, mindfulness, assertiveness and communication training, and problem-solving. QG uses push notifications to remind users to engage with the app.

### Control Intervention: Very Brief Advice

VBA is used by healthcare professionals, following the Ask, Advise, Act structure recommended by the UK government. Participants were advised to contact their local stop smoking service to access behavioral support and other forms of pharmacotherapy to help them quit smoking. Research assistants who had completed the NCSCT smoking cessation course were trained to deliver VBA, face-to-face intervention, per NCSCT guidelines (https://elearning.ncsct.co.uk). Among control group participants allocated a CO device, a mobile-app (ASH-app) was provided to enable them to view their CO results. The control group mobile-app was only used in conjunction with the CO device and contained no counselling content.

### Nicotine Replacement Therapy (NRT)

All participants were offered NRT (2mg or 4mg gum and/or 16hr or 24hr patches) free for 12 weeks, with the first two-weeks supplied at the baseline visit. Participants were also allowed to use other forms of oral NRT.

### Carbon Monoxide Monitor

A random sample of half of the participants in each treatment group were issued a carbon monoxide (CO) monitor for biochemical verification of self-reported smoking status (Smokerlyzer, coVita Inc.). CO levels were self-reported at 4-, 26- and 52-weeks post-QD. The CO devices connected via headphone/charging slot of participants’ smartphones, and were only used in conjunction with the QG and the Analyzing Smoking Habits (ASH) app, which was used to record CO measurement data. At each of the follow-up visits, participants were asked to submit a reading from their device via phone or online to validate their self-reported abstinence status. Following NCSCT guidelines, a CO reading <10ppm was considered indicative of abstinence.

### Outcome Measures

Outcome measures were collected via phone or online at baseline, 4-, 26- and 52-weeks post-quit date (post-QD). Participants received £10 to offset travel costs and were compensated for each follow-up data collection visit as follows: £10 (26 weeks) and £20 (4- and 52- week follow-ups). The primary dependent variable was continuous smoking abstinence at 26- and 52- week follow-up. Continuous smoking abstinence at each of the follow up timepoints was defined as self-reported 7-day point prevalence abstinence from cigarettes (not even a puff of smoke for the past 7 days), and expired-air CO levels of 10 ppm or lower (among the subset of individuals for whom biochemical verification was conducted). Secondary outcomes included: (1) sustained abstinence, defined as smoking no more than 5 cigarettes from the quit date to 26- and 52- week follow-up, respectively; and (2) number of quit attempts at 26- and 52-weeks post-QD.

In addition, we administered a standard demographics questionnaire, Fagerstrom Test for Nicotine Dependence (FTND) ^12^, the Smoking Abstinence Self-efficacy Questionnaire (SASEQ) ^13^, a 6-item measure used to assess changes in self-efficacy before and after the intervention within different emotional and situational contexts (24-point scale); and the Warwick-Edinburgh mental wellbeing scale (WEMWBS), a 12-item measure used to assess general positive mental health (56-point scale) ^14^. Measurements were collected via online questionnaires ^15^.

### Statistical Analysis

The sample size was determined based on the requirements linked with the primary hypothesis: evaluating differences in self-reported abstinence rates across weeks 4 through 52. Power was set at 80%, with a Type I error rate of 0.05. The estimated effect size was based upon prior literature ^16^ and factored in the anticipated attrition rate of 20%. Statistical analyses were conducted with SAS version 9.4 software (SAS Institute, Cary, NC). To determine smoking abstinence rates, both intention-to-treat (ITT) and per-protocol analyses (PP) were used. ITT analyses are considered the most conservative and are standard for psychosocial clinical trials targeting smoking cessation ^17^. Using the ITT approach, all data were analyzed, with unknown smoking status assumed to reflect continued tobacco use, resulting in conservative estimates of treatment efficacy. PP analyses excluded those for whom there were missing or unknown smoking status data and are presented herein for informational purposes, to enable comparison to ITT results.

Continuous abstinence across weeks 4, 26, and 52, based on 7-day self-reported point prevalence abstinence at each timepoint, was compared among those who received QG, relative to the control condition using mixed effects models, controlling for variables known to be associated with smoking cessation treatment outcomes, including gender, race, employment, education, and nicotine dependence severity (indicated by the FTND). Relative Risk ratios (RRs) were used to assess the outcomes for QG relative to the control group, and chi-square tests were used to test for statistical significance. Comparison of secondary outcome variables between the QG and control groups was achieved by using 2-sample *t* tests for continuous measures and chi-square tests for binary measures.

## Results

### Participants

Figure 1 displays the participant study flow. 2195 individuals were assessed for study eligibility, of which 693 were ineligible. Of the 1502 individuals eligible for inclusion, 946 failed to attend their in-person baseline visit. The remaining 556 participants eligible for inclusion in the study were randomized to study conditions (treatment n=277, control n=279). The ITT analysis included 530 participants (n=265 in each arm; 11 excluded before trial registration, and 15 for baseline protocol violations). Follow-up completion was 81.6.% at 26 weeks, and 79.4% at 52 weeks, with no difference in follow-up rates between study conditions. As shown in Table 1, participants were, on average, 41 (SD=12) years of age (range: 19 to 78). The overall sample included 291 (54.9%) men, and was predominantly Caucasian (65.5%), employed (80%), and educated, with more than two-thirds of participants reporting educational attainment beyond high school. On average, participants reported smoking 14.5 (SD=7) cigarettes per day, with an average FTND nicotine dependence score of 4 (SD=2). Most participants (85.0%, n=451) had previously made one or more quit attempts, largely by going cold turkey (48%) or using e-cigarettes (42%) and NRT (30%). There were no significant differences between study conditions in age, ethnicity, educational attainment, gender distribution, or employment status, nor in smoking behaviors, including smoking frequency, nicotine dependence severity, and previous quit attempts.

**Table 1:**
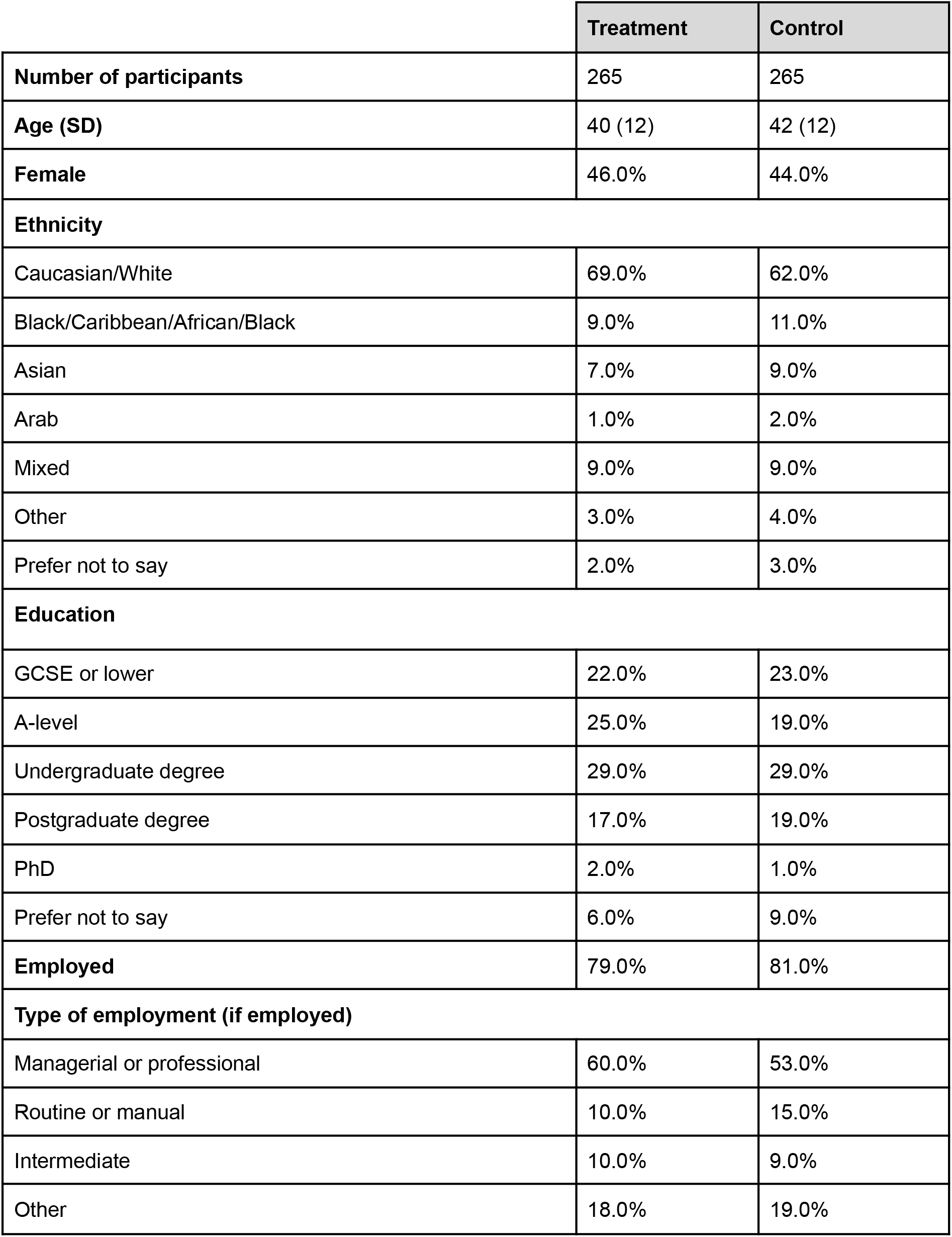
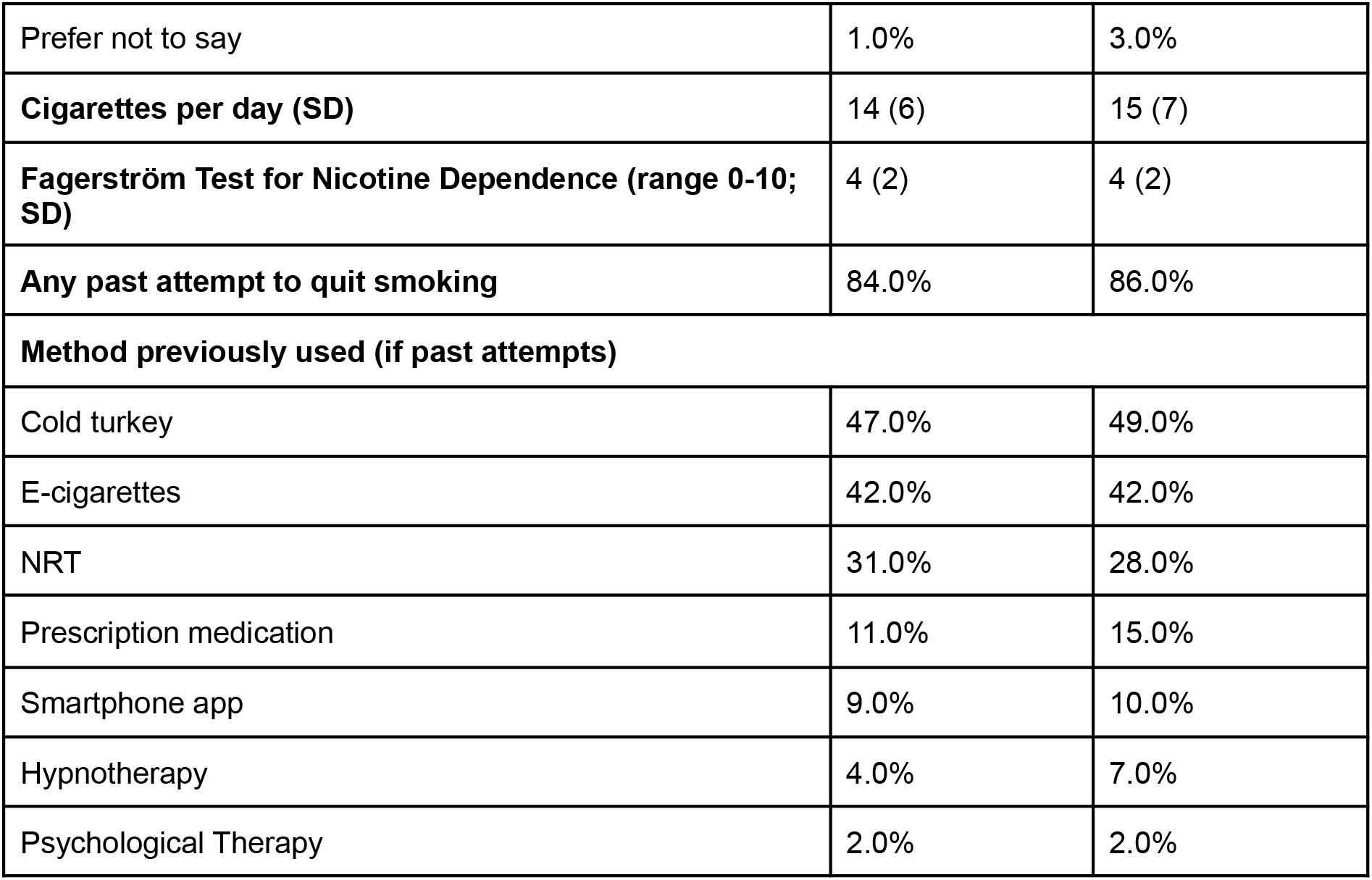
Sample characteristics.

|  | Treatment | Control |
| --- | --- | --- |
| **Number of participants** | 265 | 265 |
| **Age (SD)** | 40 (12) | 42 (12) |
| **Female** | 46.0% | 44.0% |
| **Ethnicity** |  |  |
| Caucasian/White | 69.0% | 62.0% |
| Black/Caribbean/African/Black | 9.0% | 11.0% |
| Asian | 7.0% | 9.0% |
| Arab | 1.0% | 2.0% |
| Mixed | 9.0% | 9.0% |
| Other | 3.0% | 4.0% |
| Prefer not to say | 2.0% | 3.0% |
| **Education** |  |  |
| GCSE or lower | 22.0% | 23.0% |
| A-level | 25.0% | 19.0% |
| Undergraduate degree | 29.0% | 29.0% |
| Postgraduate degree | 17.0% | 19.0% |
| PhD | 2.0% | 1.0% |
| Prefer not to say | 6.0% | 9.0% |
| **Employed** | 79.0% | 81.0% |
| **Type of employment (if employed)** |  |  |
| Managerial or professional | 60.0% | 53.0% |
| Routine or manual | 10.0% | 15.0% |
| Intermediate | 10.0% | 9.0% |
| Other | 18.0% | 19.0% |
| Prefer not to say | 1.0% | 3.0% |
| **Cigarettes per day (SD)** | 14 (6) | 15 (7) |
| **Fagerström Test for Nicotine Dependence (range 0-10; SD)** | 4 (2) | 4 (2) |
| **Any past attempt to quit smoking** | 84.0% | 86.0% |
| **Method previously used (if past attempts)** |  |  |
| Cold turkey | 47.0% | 49.0% |
| E-cigarettes | 42.0% | 42.0% |
| NRT | 31.0% | 28.0% |
| Prescription medication | 11.0% | 15.0% |
| Smartphone app | 9.0% | 10.0% |
| Hypnotherapy | 4.0% | 7.0% |
| Psychological Therapy | 2.0% | 2.0% |

**Figure 1:**
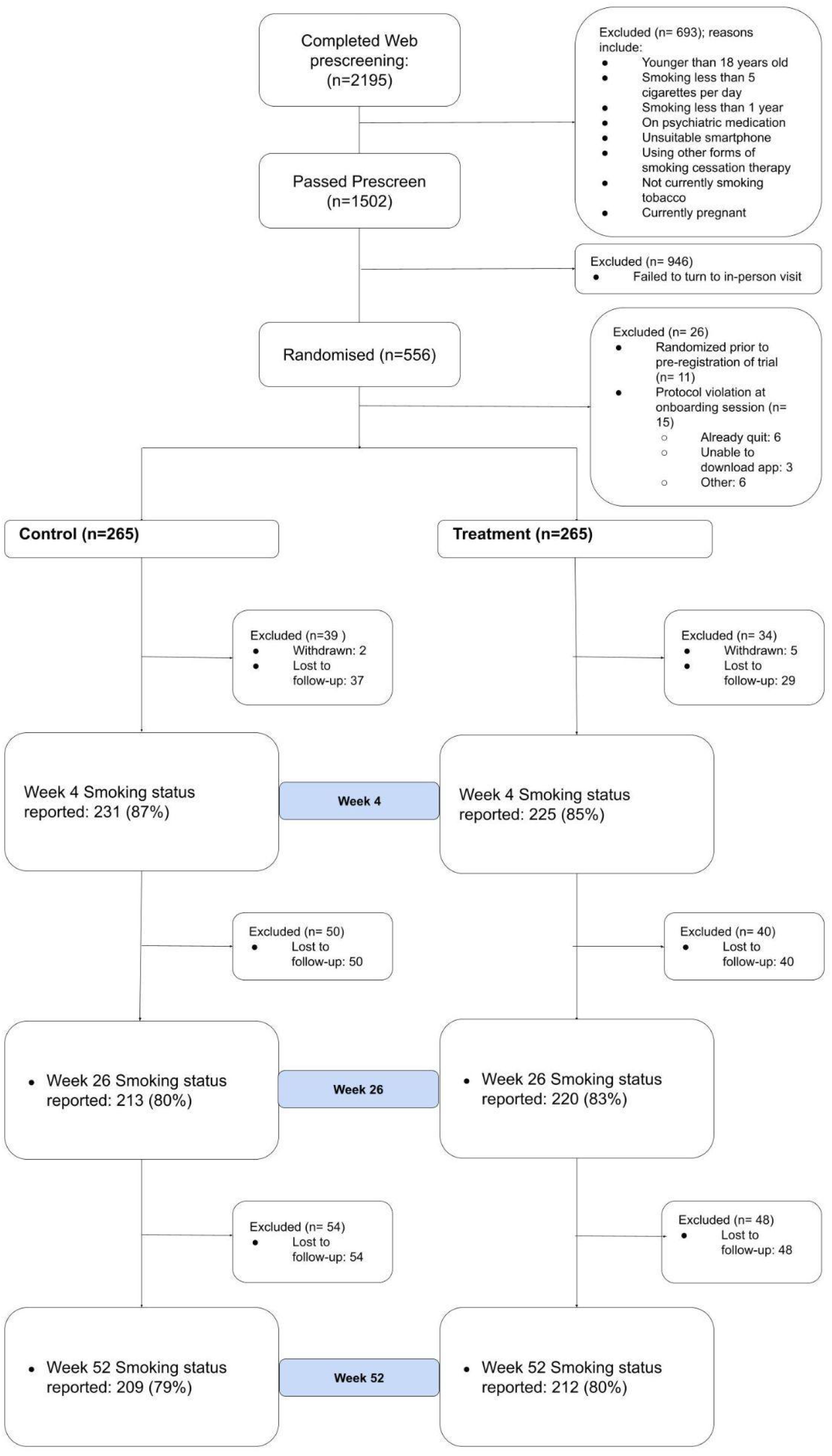
CONSORT Diagram

### Abstinence

The hypothesis that the QG condition would yield higher continuous abstinence rates across the 52-week post-quit period, compared to the control condition was supported. According to ITT analyses, after controlling for gender, demographics, and nicotine dependence severity, the odds of smoking abstinence over the 52-week course of treatment were significantly higher in the QG condition, relative to the control group (OR = 4.16, 95% CI= 2.01, 8.59; *p*<0.0001). Likewise, the QG condition produced consistently higher continuous abstinence rates relative to controls at week 26 (27.2% versus 16.6%, *p*=0.003) and week 52 (22.6% versus 13.2%, *p*=0.005) (see Table 2 and Figure 2). Moreover, relative risk ratios indicated that those who received the QG intervention and had quit successfully at 4-weeks were 70% more likely to report continuous abstinence at week 26 (95% CI = 1.22, 2.37, *p*=0.003) and 71% more likely to remain abstinent through week 52 (95% CI = 1.17, 2.50, *p*=0.005). Seven-day biochemically verified continuous abstinence rates by condition, according to both ITT and PP analyses, are shown in Table 2.

**Table 2:** Primary and Secondary Outcomes at 4-, 26, and 52 weeks Post-Quit Date

| Continuous abstinence | treatment | control | $\chi^2$ | | P-value | RR | 95% CI |
| --- | --- | --- | --- | --- | --- | --- | --- |
| 4-weeks: |  |  |  | N/A |  |  |  |
| 7-day abstinence ITT (n=530) | 118 (44.5%) | 75 (29.3%) | 13.7 |  | <.001**<br>* | 1.55 | 1.23 to 1.96 |
| 7-day abstinence PP (n=456) | 118 (52.4%) | 75 (32.5%) | 17.8 |  | <.001**<br>* | 1.62 | 1.29 to 2.0 |
| 26-weeks: |  |  |  | N/A |  |  |  |
| 7-day abstinence ITT (n=530) | 72 (27.2%) | 44 (16.6%) | 8.7 |  | 0.003** | 1.70 | 1.22 to 2.37 |
| 7-day abstinence PP (n=433) | 72 (33.8%) | 44 (20.0%) | 10.5 |  | 0.001** | 1.79 | 1.30 to 2.46 |
| (abstinent 4- and 26-weeks) |  |  |  |  |  |  |  |
| 52-weeks: |  |  |  | N/A |  |  |  |
| 7-day abstinence ITT (n=530) | 60 (22.6%) | 35 (13.2%) | 8.01 |  | 0.005** | 1.71 | 1.17 to 2.50 |
| 7-day abstinence PP (n=421) | 60 (28.7%) | 35 (16.5%) | 8.9 |  | 0.003** | 1.77 | 1.22 to 2.54 |
| (abstinent at 4-, 26- and 52-weeks) |  |  |  |  |  |  |  |
| Secondary Outcomes | treatment | control | $\chi^2$ | t test (df) | P-value | RR<sup>a</sup> | 95% CI |
| Quit attempts beyond initial QD |  |  |  | N/A |  |  |  |
| Week 4 | 68 (25.6%) | 86 (32.6%) | 2.6 |  | 0.10 | 0.79 | 0.60 to 1.03 |
| Week 26 | 94 (35.5%) | 115 (43.4%) | 3.2 |  | 0.07 | 0.82 | 0.66 to 1.01 |
| Week 52 | 104 (39.2%) | 135 (50.9%) | 6.9 |  | 0.009** | 0.77 | 0.64 to 0.93 |
| Change in<br>Self-efficacy<br>Week 4<br>Week 26<br>Week 52 | 4.2 (7.0)<br>3.8 (8.5)<br>3.3 (8.6) | 3.1<br>(6.7)<br>2.0<br>(6.9)<br>2.2<br>(7.8) | N/<br>A | 1.7<br>(527)<br>2.6<br>(506)<br>1.6<br>(524) | 0.09<br>0.01*<br>0.12 | N/A | 1.0 (-0.16 to<br>2.17)<br>1.75 (0.42 to<br>3.07)<br>1.12 (-0.28 to<br>2.52) |
| Change in<br>Mental<br>Well-being<br>Week 4<br>Week 26<br>Week 52 | 0.7 (7.0)<br>1.4 (8.2)<br>1.5 (8.4) | 0.6<br>(6.5)<br>1.0<br>(7.8)<br>0.1<br>(8.3) | N/<br>A | 0.2<br>(525)<br>0.7<br>(527)<br>1.9<br>(528) | 0.83<br>0.51<br>0.06 | N/A | 0.13 (-1.02 to<br>1.28)<br>0.45 (-0.90 to<br>1.81)<br>1.39 (-0.03 to<br>2.82) |
| 7-day point<br>prevalence<br>abstinence<br>Week 4<br>(ITT)<br>Week 4<br>(PP)<br>Week 26<br>(ITT)<br>Week 26<br>(PP)<br>Week 52<br>(ITT)<br>Week 52<br>(PP) | 118<br>(44.5%)<br>118<br>(52.4%)<br>95<br>(35.9%)<br>95<br>(44.6%)<br>92<br>(34.7%)<br>92<br>(44.0%) | 75<br>(29.3%)<br>75<br>(32.5%)<br>73<br>(27.6%)<br>73<br>(33.2%)<br>78<br>(29.4%)<br>78<br>(36.8%) | 13.<br>7<br>17.<br>8<br>4.2<br>5.9<br>1.7<br>2.3 |  | <0.001<br>***<br><0.001<br>***<br>0.03*<br>0.01*<br>0.19<br>0.13 |  | 1.55 (1.23 to<br>1.96)<br>1.62 (1.29 to<br>2.02)<br>1.32 (1.03 to<br>1.69)<br>1.38 (1.08 to<br>1.75)<br>1.20 (0.94,<br>1.54)<br>1.24 (0.99,<br>1.57) |
| Sustained<br>abstinence, <=5<br>cigs since QD<br>Week 26<br>Week 52 | 73<br>(27.5%)<br>58<br>(21.9%) | 40<br>(15.1%)<br>30<br>(11.3%) | 11.<br>5<br>9.9 | N/A | <0.001<br>***<br>0.002** | 1.82<br>1.93 | 1.29 to 2.58<br>1.29 to 2.90 |

$\chi^2$ = Chi squared test
QD = Quit Date
RR, Relative Risk ratio
ITT, Intention-to-treat analysis
PP, Per-protocol analysis
\*p<0.05
\*\*p<0.01
\*\*\*p<0.001

**Figure 2.**
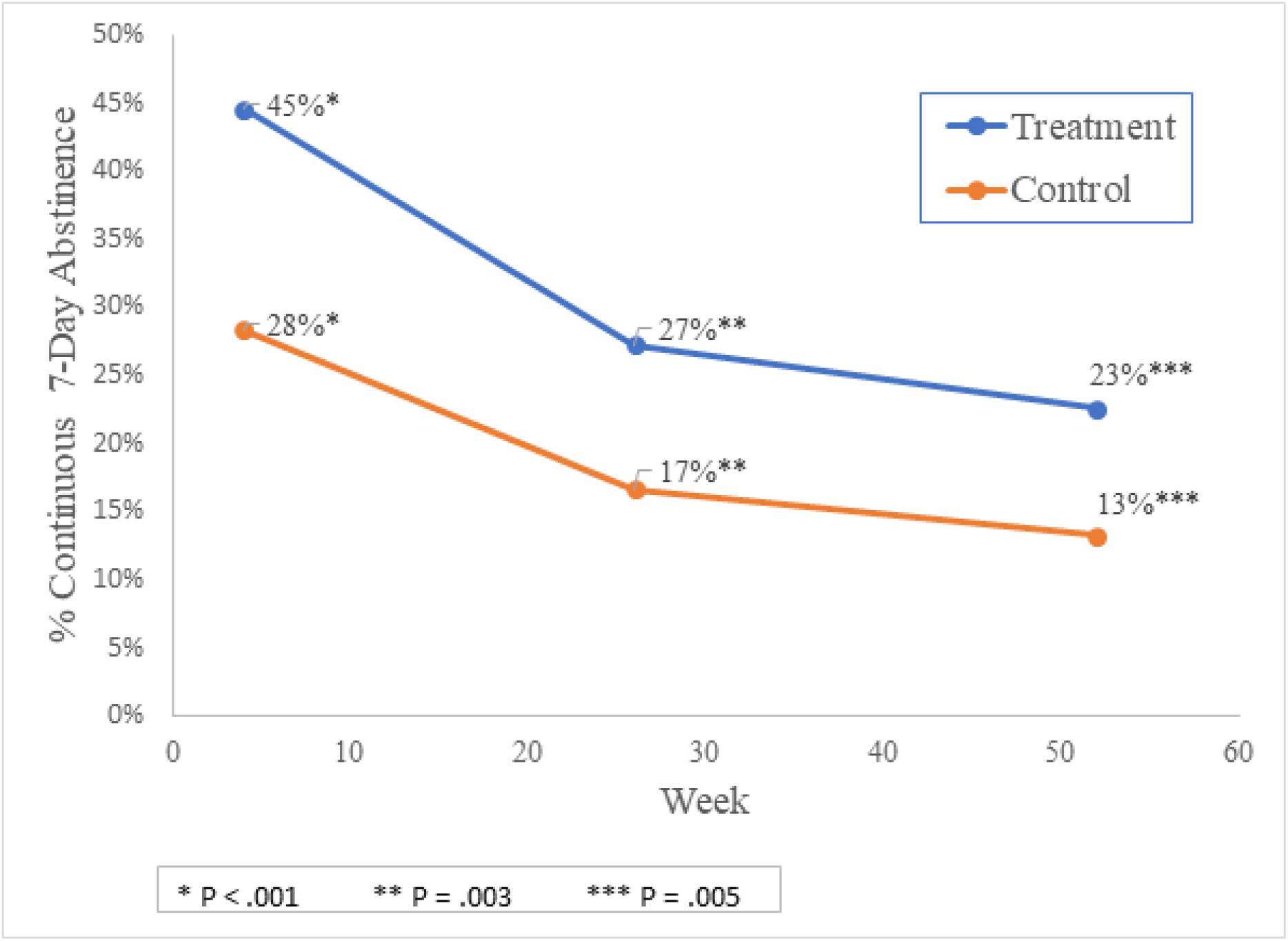
Change in Primary Outcome – 7-Day Point Prevalence Continuous Abstinence (ITT)

Among participants who were assigned a CO monitor, 97.1% (n=134) in the QG condition and 97.9% (n=139) in the control group provided a baseline CO reading. Among those who subsequently reported abstinence, at 4-weeks post-QD the expired-air CO levels of 10 ppm or lower corresponded with participant self-report for nearly all (96.4%; n= 80) participants. At 26- and 52-weeks, self-reported abstinence was biochemically verified among 98.4% and 95.5% of participants, respectively.

The QG condition produced consistently higher continuous abstinence rates at weeks 4, 26, and 52, relative to the control group (p<0.01 at each assessment point), with the exception of self-reported 7-day point prevalence of abstinence at 52 weeks (p>0.05) (See Table 3). QG participants evidenced higher rates of sustained abstinence compared with those who received the control intervention, both at 26-weeks (RR=1.82, 95% CI, 1.29 -2.58 ;27.5% vs 15.1% quit rate) and at 52-weeks (RR= 1.93, 95% CI 1.30, 2.91; 21.9% vs 11.3% quit rate).

As shown in Table 2, for the first 26-weeks of the study, there was no difference between treatment conditions in additional quit attempts after the initial quit date (QG = 35% versus Control = 43%, p>0.05). At 52-weeks, however, participants in the control group were significantly more likely to have reported additional quit attempts (RR= 0.77, 95% CI 0.64 - 0.93).

The hypothesis that those who were assigned to the QG condition would demonstrate greater changes in putative mechanisms of action of CBT relative to those assigned to the control intervention, including self-efficacy and mental well-being, was partially supported. Though differences in self-efficacy for managing smoking urges were not observed at week 4 (*p*=0.09), a difference emerged at week 26, with larger increases in self-efficacy observed among those who received the QG intervention, *t*(506) = 2.6, p=0.01. However, at 52-weeks, that effect had diminished (*p*=0.12). No differences between groups were detected in regards to participant mental well-being at any of the 3 timepoints.

## Discussion

The purpose of this study was to evaluate QG, an extended CBT-based digital therapeutic intervention combining pharmacotherapy and behavioral treatment for smoking cessation. Based on the well accepted chronic disease model of addiction ^18^, the 52-week QG treatment program produced continuous abstinence rates at 26 and 52 weeks post-QD that are substantially higher than those typically reported in the literature. QG participants who effectively quit smoking 4 weeks post-QD were 1.70 times more likely to remain abstinent relative to control group participants at week 26 (27.2% abstinent versus 16.6% abstinent), and 1.71 times more likely to remain abstinent at week 52 (22.6% abstinent versus 13.2% abstinent).

Importantly, these findings extend the preliminary outcomes previously reported, establishing not only the short-term efficacy of the QG intervention in producing high abstinence rates (44.5% at 4 weeks post-QD, according to ITT analyses), but also its superiority in helping smokers achieve repeated abstinence over the course of one year. The intervention approach used in this study differs from others reported in the literature in that it combines psychosocial and pharmacological modalities, and concurrently provides access to the psychosocial component over an extended time period, regardless of participants’ abstinence status over that timeframe. Thus, the current study advances the evidence base for extended, digital health intervention models for smoking cessation. Although point prevalence abstinence rates have been reported across various RCTs of digital health interventions, according to a recent Cochrane review of smartphone and app-based smoking cessation interventions, 6-month abstinence rates ranged from 4% to 18% ^19^, suggesting that the extended, multi-modality approach inherent to the QG program is an advance compared with existing digital interventions for individuals with tobacco addiction. Moreover, longer term quit rates reported in the literature typically refer to the proportion of individuals abstinent at a given point in time, rather than capturing continuous abstinence across multiple follow-ups, a more stringent measurement of favorable treatment outcomes. Thus, the 26-week point prevalence abstinence rate of 35.9% observed among QG participants not only exceeds long term quit rates reported for other digital smoking cessation interventions, such as iCanQuit, an ACT-based smoking cessation application (29.6% quit rate at 6 months post-treatment) ^20^, and Craving to Quit, a mobile app based mindfulness training program (11% abstinence rate at 6 months post-baseline) ^21^, but our primary outcome of continuous abstinence at 6 and 12 months (27.2% and 22.6%, respectively) far exceeds those reported for various digital programs, including iCanQuit (prolonged abstinence rate of 13.8% at 12 months post-treatment) ^20^, Happy Quit, a text messaging based intervention (6.5% at 6 months post-treatment) ^22^, and is comparable to Pivot’s 23.8% (ITT) continuous abstinence at 7 months post-enrollment ^23^.

While there is limited literature surrounding digital therapeutic interventions for smoking cessation beyond 6 months post-treatment, the continuous abstinence rate observed at 52 week follow-up in the present study is substantially higher than that observed in the handful of studies of digital smoking cessation treatments with long-term follow up measurements, in which prolonged abstinence rates are below 10% by 12 months ^20^.

Methodological limitations of extant studies, including the use of single-arm designs, coupled with the absence of biochemical verification of self-report data, pose challenges to interpretation and generalizability of prior findings. To overcome these limitations, the present study employed a 2-arm parallel-group RCT design with biochemical verification. Despite verifying the CO of a subset of participants, the high correspondence between CO readings in this study and self-reported abstinence is consistent with conclusions of a review by the Society for Research on Nicotine and Tobacco Subcommittee on Biochemical Verification ^24^, indicating that biochemical validation is not always necessary in smoking cessation studies, because levels of misrepresentation are generally low (0–8.8%).

There were no significant group differences in quit-attempts at 4- and 26-weeks; however, at 52-weeks, despite having lower quit rates, control group participants were significantly more likely to have reported additional quit attempts beyond their initial QD than those in the treatment group, suggesting that they continued to attempt, albeit less successfully, to stop smoking. Focusing on re-engaging individuals who initially failed to quit and increasing their motivation to reinitiate quit attempts will likely improve future success rates of QG.

Self-efficacy is an important process variable underlying successful smoking abstinence outcomes ^25,26^. Improvement in self-efficacy was greater among those who received QG, relative to controls, an effect that emerged at 26-weeks post-QD. These findings replicate and extend the increase in reported quit-confidence and self-efficacy observed in Webb et al’s (2020) preliminary outcome findings, indicating that digital CBT based interventions can affect similar psychological process variables to face-to-face treatments ^10^. These differences in self-efficacy diminished at 52-weeks post-QD, suggesting that understanding and addressing barriers to maintaining confidence is a potential target for future intervention refinement. Though overall mental well-being improved generally among participants, there were no group differences in the magnitude of improvement, suggesting that further research is needed to build upon our understanding of how digital interventions can positively target mental wellness among smokers. Nevertheless, given that those receiving psychiatric medication were excluded from participation in this study, the absence of effects on mental well-being may be attributable to a restricted range of mental well-being in the study sample.

### Strengths and Limitations

There are several strengths of this study, including a large sample size, the randomized controlled design, the use of remote biochemical verification and high correspondence between self-reported smoking status and CO measurements, the high participant retention rate and long-term follow-ups.

This study also has some limitations. First, QG was not compared to another digital intervention. VBA was used as the control intervention due to its frequent use as the first-line intervention for smoking cessation in the UK, and for the participants not allocated a CO device, no mobile-app was provided. To control fully for time and attention, future studies should consider incorporating a digital intervention control condition.

Second, this study relied largely on self-reported smoking abstinence status, which may have been exaggerated. To mitigate this, we verified self-reported outcomes using a CO measurement device among 50% of participants in each study condition. Despite the limited biochemical verification data, the consistently high level of agreement between the CO readings and self-reported abstinence across all study timepoints suggests that, in the context of this study, self-report is a reliable indicator for true smoking abstinence.

Third, researchers were unblinded to participant group allocation. Finally, there are some limitations to the generalizability of the study findings, in light of the characteristics of the study sample. Participants with serious health conditions and/or who were using psychiatric medication were excluded. In light of the high rates of psychiatric comorbidity among smokers, replication and extension of the study to include those with co-occurring mental health conditions appears warranted. Further, given that the study sample comprises a largely urban population, the long term efficacy of a digital health intervention such as QG among rural groups of smokers, who could benefit tremendously from the accessibility of this approach, remains unknown.

## Conclusion

QG, a DTI utilizing an extended care model combining NRT with evidence-based psychosocial treatment for smoking cessation, was highly effective in achieving continuous smoking cessation at 4-, 26- and 52-weeks compared to the control group using VBA.

Still, opportunities exist to improve psychological outcomes such as mental well-being and self-efficacy through future intervention refinement. Studies of the effectiveness of QG among more diverse populations of smokers are a promising area for future research.

## Data Availability

Available upon request

## Acknowledgements

The authors thank the NHS primary care practices in the London districts of Barnet, Camden, Ealing, Hammersmith and Fulham, Islington, Kilburn and North Kensington for their help in supporting participant recruitment. AM is supported by the NIHR NIHR Applied Research Collaboration NW London. The views expressed in this publication are those of the authors and not necessarily those of the NIHR or the Department of Health and Social Care.

